# OUTSIDE 2: Outdoor swimming as a nature-based intervention, offered in addition to usual care, compared to usual care alone, in people with depression: A study protocol for a randomised controlled trial and economic evaluation

**DOI:** 10.64898/2026.03.17.26348590

**Authors:** Heather Massey, Hannah Denton, Anna-Marie Bibby-Jones, Stephen Bremner, Mara Violato, Amy Burlingham, Rebecca Cunningham, Sabrina Hasnaoui, Mark Harper, Sam Robertson, Sandy Ciccognani, Kay Aranda, Amy Arbon, Gail Murphy, Chloe Bruce, Debbie Lambert, Clara Strauss

## Abstract

**Background:** Major depression affects at least 10% of adults, yet evidence-based treatments have modest clinical effectiveness and acceptability. In recent years, outdoor swimming has grown increasingly popular and emerging quantitative and qualitative evidence suggests potential as an intervention for depression, however, there is yet to be a full-scale randomised controlled trial (RCT). This is a protocol to assess the safety and test the clinical- and cost-effectiveness of an 8-session outdoor swimming course (in addition to usual care) on depression symptom severity in adults experiencing major depression, in comparison to usual care alone.

**Methods:** This study is a pragmatic, parallel group, superiority RCT with 1:1 allocation comparing the outdoor swimming intervention (in addition to treatment as usual) with treatment as usual, aiming to recruit 480 adult participants meeting diagnostic criteria for major depression. Recruitment will take place across 21 sites with blind post treatment and follow up assessments. The primary outcome is depression symptom severity at T1 post-intervention, 12 weeks post-randomisation (the primary end point) using the Patient Health Questionnaire 9 (PHQ-9). Secondary clinical outcomes are anxiety (Generalised Anxiety Disorder 7 at T1 and T2 [38 weeks post-randomisation] and PHQ-9 at T2), mindfulness is measured as a potential mechanism at all timepoints (Five Facet Mindfulness Questionnaire 15). Health economic measures at all time points are: EQ-5D-5L, Recovering Quality of Life (ReQoL), Client Service Receipt Inventory and the Productivity Cost Questionnaire. A qualitative study will examine the experience of participants during and after the swim course.

**Discussion:** If the 8-session outdoor swim course is safe, clinically- and cost-effective, findings will support national implementation, offering an evidence-based intervention to those affected by depression, while potentially reducing healthcare costs. Furthermore, this may pave the way for other outdoor activities to support people with poor mental health to be developed and evaluated as interventions.

**Trial registration:** Controlled trial registration number is ISRCTN registration number 24759023. Registered on 21 February 2024 (https://www.isrctn.com/ISRCTN24759023).

**Administrative information:** Note: Numbers enclosed in braces within this protocol correspond to SPIRIT checklist item numbers. The sequence of items has been adjusted to group related content together. (https://www.consort-spirit.org/).

|  |  |
| --- | --- |
| Title {1} | OUTSIDE 2: Outdoor swimming as a nature-based intervention, offered in addition to usual care, compared to usual care alone, in people with depression: A study protocol for a randomised controlled trial and economic evaluation |
| Trial registration {4}. | ISRCTN 24759023 date of registration: February 21, 2024 (prospectively registered). |
| Protocol version {2} | Version 11.0 08/07/2025 |
| Funding {7a} | NIHR PRP (ST-01-51) reference number 203462 |
| Name and contact information for the trial sponsor {3b} | Research and Development Department<br>Sussex Partnership NHS Foundation Trust<br>Sussex Education Centre<br>Nevill Avenue<br>Hove<br>BN3 7HZ |
| Role of sponsor {3c} | The sponsor is accountable for research support and funding, has contributed to the study's design, and retains final decision making authority regarding the publication of this report. |

**Plain Language Summary:** 

**Background:** Depression is very common,with at least 1 in 10 people having an episode of depression during their lifetime. Many people believe that outdoor swimming can improve depression. There is some research that suggests outdoor swimming is helpful for depression, but we do not yet have enough evidence to be sure. OUTSIDE 2 is a large research study that aims to find out whether outdoor swimming can help people with depression.

**Aim:** The aim of this study is to find out if taking part in an 8-session outdoor swimming course, along with usual care, is safe, helps improve depression, and is good value for money. We will compare people who do the swimming course (alongside their usual care) with people who only receive their usual care. In our study we will also talk to participants about their experiences. We want to understand how the outdoor swimming course affects their depression, and how the activity itself might help them feel better.

**Describe your research plan, including:** People who are interested in taking part in the study can visit our website (outside2.co.uk) to read more information and sign up. Once someone agrees to join, they will be randomly placed into one of two groups available at their location to keep the study fair. One group will take part in eight one-hour outdoor swimming sessions in a small group. They will continue with their usual care for depression, which may include talking therapies, antidepressant medication, or community activities. The other group will continue with usual care only during the study. After the study ends, they will be offered the same outdoor swimming course, so no one misses out.

The swimming sessions will be led by experienced swimming coaches who will help participants build confidence in the water and learn important water safety skills. During the study, all participants will complete questionnaires about their symptoms of depression, overall mental health, and use of health services. They will do these before they are put in a group, right after the swimming course or usual care period and six months later.

Participants in the swimming group will also be asked to keep a diary about their experiences during the course. Researchers may visit some sessions to ask participants about how they are finding the swimming and how it affects their mood. Swim coaches will record attendance at each session and describe what activities were included. This helps the research team understand exactly what took place during each class.

**Knowledge mobilisation:** We want to make sure that the information we learn from this study reaches many different people, not just scientists. To do this, we will share our findings in several ways. We will create a Podcast mini-series, an animated video as well as research papers. Using these different methods helps us share our findings with many groups, including adults in the community, mental health professionals, people working in the swimming industry, scientists, and policymakers. By sharing the results in several ways, more people can understand if and how outdoor swimming might support recovery from depression and mental health more broadly, and it increases the chance that the study will make a real difference, whatever it finds.

## Introduction

### Background and rationale {9a, 9b}

Depression affects approximately one in ten adults over the course of their lives^1^. In England, the economic impact of mental ill health was estimated at £105 billion in 2016^2^, and between 2023 and 2024, nearly 89 million prescriptions for antidepressants were issued^3^. Among individuals diagnosed with depression, 10–20% experience a chronic course, with symptoms persisting for more than two years, while approximately 50% are at risk of recurrent episodes throughout their lifetime^4^.

Given the prevalence and complexity of depression, effective and sustainable treatment strategies are urgently needed. Current guidelines from the National Institute for Health and Care Excellence (NICE) recommend a range of evidence-based interventions, including psychological therapies, pharmacological treatments, and structured exercise programmes^5^. However, despite extensive research, the effectiveness of these interventions remains modest^6,7^, and adherence to both medication and psychological therapies is often limited^8,9^. Emerging evidence indicates that engaging in physical activity at levels recommended by the World Health Organization (WHO)^10^ of 150 minutes per week confers protective effects against elevated all-cause mortality in individuals with major depressive disorder^11^. The therapeutic efficacy of exercise in managing depression is well-established^12,13^. However, population-level adherence to physical activity guidelines remains insufficient in the UK, with only 70% of men and 59% of women meeting the recommended thresholds of 150 minutes of moderate-intensity or 75 minutes of vigorous-intensity activity per week^14^.

These limitations have prompted growing interest in alternative approaches to mental health care. One such approach involves exposure to natural environments, particularly green (areas of grass, trees or vegetation) and blue spaces (areas that feature predominately water), which has been increasingly associated with improved mental health outcomes^15–17^. However, factors such as accessibility, personal safety, and preference may influence participation. A diverse range of activities, including nature-based options, will be necessary to ensure individuals can identify and engage with forms of physical activity that align with their personal preferences, interests, and needs.

Outdoor swimming, a form of blue exercise, has seen a rise in popularity^18^. Participants frequently report enhanced wellbeing, a deeper connection to nature, and significant mental health benefits^19–23^. Despite these promising anecdotal and preliminary findings, no full-scale randomised controlled trial (RCT) has yet been conducted to evaluate its clinical or cost-effectiveness. Moreover, outdoor swimming carries inherent risks^24–26^, and access to blue spaces is not equitable^27,28^. Barriers include limited mobility^29^, socioeconomic disadvantage^30,31^, and for people who have not learned to swim may contribute to feelings of exclusion^32,33^.

To assess the feasibility of outdoor swimming as a therapeutic intervention for depression, we previously conducted a feasibility RCT involving an outdoor swimming programme delivered in 8 sessions by qualified open water coaches. The intervention was offered alongside usual care and compared to usual care alone in adults with mild to moderate depression^34,35^. Feasibility outcomes were met with high recruitment (99%) and retention rates (84%). Although the study was not intended to detect statistically significant differences, between-group comparisons were encouraging.

The Patient Health Questionnaire-9 (PHQ-9)^36^, proposed as the primary outcome measure of depression symptom severity, demonstrated sensitivity to change, with a medium-to-large effect size favouring the intervention over treatment-as-usual (TAU) at post-intervention (Cohen’s d = 0.71, 95% CI [1.88-5.52])^35^. Health economic outcomes were also encouraging, with indications of improved health-related quality of life in the intervention group in comparison to TAU.

An embedded qualitative study explored barriers and facilitators to participation, identifying five key themes: Accessibility, Belonging, Facing Challenges with Support, Benefiting and Enjoyment, and Clarity of Information. These insights informed several recommendations for future research, including reducing assessment burden, tailoring courses for underserved populations, improving clarity of study materials, and enhancing accessibility of participant information^35,37^.

The current study builds on this foundation by conducting a full-scale RCT to evaluate the safety, clinical- and cost-effectiveness of the same 8-session introductory outdoor swimming intervention in adults experiencing a current episode of major depression. Alongside this we will explore the experience participants have of being part of the outdoor swimming course. If demonstrated as effective, this intervention could be scaled nationally, improving access to nature-based mental health support and potentially benefiting individuals, the National Health Service (NHS), and the wider economy.

### Objectives {10}

The objectives of this research study are to assess the safety and test the clinical- and cost-effectiveness of an 8-session outdoor swim course (in addition to usual care) on depression symptom severity in adults experiencing a current episode of major depression, in comparison to usual care alone. The proposed study will assess the following hypotheses:

### For adults experiencing an episode of major depression

#### Hypothesis 1

The outdoor swimming course plus usual care, compared to usual care alone, will result in larger reductions in depressive symptom severity from baseline (Time point 0, T0) to post intervention 12 weeks following randomisation (primary outcome/endpoint, Time point 1, T1).

Secondary Hypotheses are that the outdoor swimming course plus usual care, compared to usual care alone will:

#### Hypothesis 2

result in larger reductions in depressive symptom severity from T0 to 38 weeks following randomisation (follow-up, Time point 2, T2).

#### Hypothesis 3

result in larger reductions in anxiety symptom severity from T0 to T1 and from T2.

#### Hypothesis 4

result in larger improvements in mindfulness from T0 to T1 and from T2.

#### Hypothesis 5

be cost-effective at T2.

An embedded qualitative study will explore participants’ experiences of the outdoor swimming course, with a particular focus on how the physical activity of outdoor swimming interacts with their experience of depression.

### Trial design {12}

<u>OUT</u>door <u>S</u>wimming Intervention for <u>DE</u>pression (OUTSIDE 2) is a parallel-group, pragmatic superiority RCT with 1:1 allocation to an outdoor swimming course plus usual care or usual care alone. Depression symptom severity using the PHQ-9 is the primary outcome and will be collected at three time points: baseline (T0) a maximum of four weeks before the first session of the swim course; post intervention (T1), at 12 weeks post randomisation: and follow up (T2), at 38 weeks post randomisation. The primary clinical endpoint is T1. The main assessment point for the economic evaluation is T2 with the primary health economic outcome being incremental cost per Quality Adjusted Life Year (QALY) gained.

The embedded qualitative component will focus on the experience of being on the swim course and use diaries and interviews, during and after completion of the course, to collect data.

## Methods: Participants, interventions and outcomes

### Patients and Public Involvement (PPI) {11}

Patient and public involvement (PPI) was used extensively during the design and delivery of the feasibility OUTSIDE 1^35,37^, and it will again play a central role in supporting the delivery of OUTSIDE 2. We have supported a group of people with lived experience of depression or other mental health difficulties to help us design the website and participant information materials, ensuring that everyone has the information they need to decide whether they would like to take part in the study.

PPI will be embedded throughout the development and delivery of OUTSIDE 2. The study will be supported by a Lived Experience Advisory Panel (LEAP), made up of members of the public with direct experience of depression, outdoor swimming, or mental health challenges, and their contributions will help shape the study’s priorities. The LEAP will meet regularly and will provide ongoing advice throughout the study. Early involvement of LEAP members will help the research team understand the study’s relevance from a public perspective and ensure that the work remains grounded in the experiences of those it aims to support.

The research questions will be developed and refined through regular discussions with the LEAP. During these meetings, the research team will present initial ideas and invite LEAP members to share their perspectives. Their input will guide decisions about participant engagement, recruitment approaches, and potential barriers to inclusion. These early discussions will also help the team consider issues related to equality, diversity and inclusion (EDI), particularly in relation to underserved communities.

LEAP members will contribute to study design by providing feedback on the proposed research methods, including the outdoor swimming course, qualitative diaries, interviews, and participant⍰facing materials. While some study elements such as clinical outcome measures will be selected for scientific reasons, LEAP members will help the team acknowledge limitations and interpret findings in ways that are meaningful to the people the study intends to benefit. Because the study will recruit participants directly, LEAP members will not be involved in participant recruitment, but they will advise on how to make the study more accessible and welcoming.

Patients and the public will also contribute to the development of the study’s dissemination plans. LEAP members will advise on how findings should be communicated to non⍰academic audiences and help identify groups and communities who may be particularly interested in the results. They will provide feedback on public⍰facing outputs, and written summaries, ensuring that these materials are clear, accessible and reflective of lived experience. This will help ensure that the study’s findings reach beyond academic audiences and support wider understanding of how outdoor swimming as part of a group led by an experienced coach may help people living with depression.

### Study setting {13}

The study will recruit from 21 locations across England, with good geographical and sociodemographic coverage. The swim courses will take place at a variety of types of outdoor swim sites including the sea, rivers, lakes and lidos (outdoor swimming pools). The type of water, and swimming coach qualifications will determine what level of swimming experience is required of participants to join the courses and therefore the trial (see below).

### Eligibility criteria {14a, 14b}

Decisions about inclusions and exclusions will be based on medical information provided by participants and reviewed by the trial medical team, but further information or tests ordered via the participant’s General Practitioner may be required if indicated.

Inclusion criteria are:

1. Participants give fully informed consent to participate;
2. Clinically important symptoms of depression, as determined by the Patient Health Questionnaire 9 (PHQ-9) score of 8 and above
3. Meeting DSM 5 criteria for a current Major Depressive Episode on the Mini International Neuropsychiatric Interview [MINI 7.0.2]^38^ (the MINI 7.0.2 is a short structured diagnostic interview with screening and full versions to assess for DSM-5 and ICD-10 psychiatric disorders);
4. Self-reported swimming competence, being able to swim a minimum distance in a heated indoor pool. This differed by the location participants will attend and is a requirement to be able to keep participants safe in these environments; for sea locations (50 m, 2 lengths of a normal swimming pool) and lake locations (25 m, 1 length of a normal swimming pool), no swimming ability is required for outdoor unheated swimming pools (lidos) in accordance with our previous feasibility trial^34,35^;
5. Adult aged 18 years or older;
6. Able to understand spoken instructions in English or in a language spoken by the swim coaches that are recruited.

Exclusion criteria are:

1. High risk of suicide (determined by asking the person reporting: (1) in the past month having taken active steps with the intention of dying, (2) greater than 10% risk of acting on suicidal thoughts in the next few days (3) or 50% or greater risk of acting on suicidal thoughts in the next 3 months, or (4) for those reporting 1-49% risk of acting on suicidal thoughts in the next 3 months, risk of acting on thoughts deemed high following review by a study psychiatrist);
2. Evidence of a current Psychotic Disorder, or experiencing acute psychotic symptoms in the last 6 months (ascertained by the MINI 7.0.2, further assessed by the study psychiatry team where necessary); Self-reported using a health history questionnaire (see Appendix 1), followed up with the medical team if necessary, with either further discussion or further investigation:
3. History of serious cardiac abnormalities;
4. Respiratory conditions exacerbated by cold, for example self-reported poorly controlled exercise-induced asthma;
5. Cold-water urticaria;
6. Participants with a BMI of 17 or under;
7. Moderate to severe learning disability;
8. History of non-freezing cold injuries, if not mitigated (wearing boots/gloves). Or,
9. Currently participating in another interventional research trial

### Who will receive informed consent? {32a}

Upon receiving an expression of interest via the study website, prospective participants will be sent a participant information sheet and an online consent form. These documents will be provided at least 24 hours prior to a scheduled eligibility assessment, which will be conducted by a trained research assistant (RA) via telephone or video conference, with an in-person option offered if required. During the eligibility assessment, inclusion and exclusion criteria will be reviewed, and participants will have the opportunity to ask questions about the study to support informed decision-making.

### Additional consent provisions for collection and use of participant data and biological specimens {32b}

During the consent process, participants will be informed that their data will be retained even if they choose to withdraw from the trial. They will also be asked to provide explicit permission for the research team to share relevant data with representatives of participating NHS Trusts and, where appropriate, regulatory authorities. This trial will not involve the collection or storage of biological samples.

### Interventions

A two-arm RCT will be conducted to (1) isolate the effects of the outdoor swimming intervention course from those of usual care, and (2) assess the cost-effectiveness of the intervention in comparison to usual care.

### Intervention description {15a}

#### Intervention

The structure of the swimming course will replicate the approach used in the OUTSIDE 1 feasibility study^34,35^. Participants allocated to the intervention arm will be invited to attend either an online or in-person introductory session, lasting approximately one hour, led by the outdoor swimming coaches. This session will take place prior to the commencement of the 8-week course and will provide an opportunity for participants to meet the coaches and ask questions about the programme. All coaches will either have prior experience supporting individuals with mental health challenges or will be trained in mental health first aid. In addition, coaches will receive study-specific training before the intervention begins, covering research procedures and session content.

The outdoor swimming intervention will consist of weekly sessions, each lasting approximately one hour, delivered over an 8-week period. Courses will commence within four weeks of randomisation and will include between 5 and 11 participants per group. The sessions will prioritise safe and enjoyable engagement with the water, rather than focusing on swimming technique or physical fitness. Participants will be encouraged to continue with their usual care, including any prescribed medications or psychological therapies, throughout the intervention period. They will also be supported to identify existing swimming groups or form new ones to facilitate continued safe participation following the course. Following the course, outcome measures for clinical and economic evaluation will be collected at 12 weeks post-randomisation (T1), with a follow-up survey administered at 38 weeks post-randomisation (T2).

Outdoor swimming carries inherent risks, to mitigate these risks, the intervention will be delivered in line with strict eligibility criteria that exclude individuals with physical conditions contraindicated for cold water immersion in line with Tipton et al^25^. All sessions will be led by qualified and experienced open-water swimming coaches and appropriately qualified lifeguards. The intervention will be delivered during the UK spring and summer months (April to September 2024 and 2025), when water temperatures are typically at their highest (estimated between 14°C and 24°C).

Each swim session will include a pre-swim briefing focused on safety, a group activity or shared experience, and a period of self-directed time in the water. While the core structure will be consistent, session content will be adapted to suit the location, weather conditions, and participants’ swimming confidence. Coaches will complete a weekly checklist to record attendance and session activities.

Participants will continue to access health and social care services as usual, with no restrictions imposed by trial participation.

#### Comparator: Usual care control

The usual care arm will serve as the control condition. Participants allocated to this arm will continue to access healthcare services without restriction. During the trial period, they may be prescribed medications, placed on waiting lists, engaged in psychological therapies, or involved in social prescribing activities. Engagement with other treatments or interventions will vary, and participants will be asked to record any additional therapies or services they access throughout the study. Clinical and health economic outcome data will be collected at the same time points as the intervention group, using online surveys or with support from RAs where needed.

Participants in this arm will be given the chance to join an introductory outdoor swimming course following the trial.

### Criteria for discontinuing or modifying allocated interventions {15b}

Participants will be clearly informed that they may withdraw from the intervention and/or the study at any time, without any impact on their access to usual health or social care services. Should any aspect of the study, including the intervention, cause distress or discomfort, participants will be supported in making an informed decision about continuation. The intervention may also be discontinued if, in consultation with at least one of the Chief Investigators and the participant, the swim coach or medical team determines that continued participation poses a risk to the participant’s mental or physical health. In such cases, participants may choose to remain in the study and will be included in the intention-to-treat analysis.

### Strategies to improve adherence to interventions {15c}

To support adherence to the trial, participants randomised to the usual care control arm will be offered the intervention at the end of the study, as a thank you for their participation. In addition, travel reimbursement for attendance at the intervention swim course will be provided.

Adherence to the intervention will be monitored through an attendance register kept by the swim coach and reported after each of the intervention courses sessions. Swim coaches will be encouraged to reach out to people who do not attend to welcome them to the next session.

### Relevant concomitant care permitted or prohibited during the trial {15d}

Participants in both trial arms will be encouraged to continue engaging with their existing health and social care services throughout the study period. To systematically monitor service utilisation, the research team will employ an adapted version of the Client Service Receipt Inventory (CSRI)^39^, enabling the collection of detailed data on health and social care use across a range of services. This approach will support a comprehensive evaluation of resource use and inform the health economic analysis.

### Provisions for post-trial care {34}

No provision for post-trial care will be made within the study. Participants will remain under the care of their usual health and social care providers following the conclusion of the trial.

### Outcomes {16}

#### Screening measures

- Mini International Neuropsychiatric Interview [MINI 7.0.2]^38^.

The MINI 7.0.2 is a short structured diagnostic interview with screening and full versions to assess for DSM-5 and ICD-10 psychiatric disorders). Current symptoms of depression, psychosis, and suicide risk will be screened using the MINI-screen, with full MINI modules administered as needed to more comprehensively assess major depression and psychosis.See exclusion criteria for details on how high risk of suicide was determined.

#### Demographic measures

- Information About You survey: Age, Sex, Gender, Whether Transgender, Employment, Sexual Orientation, Marital Status, Birth Country, First Language, Ethnicity, Education, Disability Status, Number of children under/over 18 years, Number of Adults cared for, Religion or belief, Index of Multiple Deprivation, Housing status, Income, Level of swimming ability, Previous experience of swimming, Previous treatment for depression.

#### Clinical measures

- Primary Outcome: Patient Health Questionnaire-9 (PHQ-9)^36^ is a nine-item self-report measure of depression severity with established sensitivity and specificity. Items are rated on a four-point scale. Scores of 5–9 indicate mild depression, 10–14 moderate, 15–19 moderately severe, and 20 or above severe symptoms^36^. The measure is included because the intervention is hypothesised to reduce depressive symptoms, which may change over time. Assessments will occur at baseline (T0), 12 weeks post-randomisation (T1), and 38 weeks post-randomisation (T2). The primary outcome will be the change in PHQ-9 scores between baseline (T0) and 38 weeks post-randomisation (T2).
- Secondary (Clinical) Outcomes: Generalised anxiety will be assessed using the Generalised Anxiety Disorder-7 (GAD-7) ^40^, a seven-item measure rated on a four-point scale with robust psychometric properties ^41^. Assessments will occur at T0, T1, and T2.
- Secondary (Mechanism) Outcomes: Mindfulness will be evaluated using the 15-item version of the Five Facet Mindfulness Questionnaire, which has shown excellent psychometric reliability and validity^42^. Moderate between-group mean differences for mindfulness were identified in OUTSIDE 1^34,35^, supporting its inclusion in this survey. Evaluation time points will be at T0, T1, and T2.

#### Health economic measures

We will collect the following health economics measures at time points T0, T1 and T2:

- The EQ-5D-5L^43^ will be used to assess generic health-related quality of life (HRQoL). The EQ-5D-5L is a preference-based instrument commonly applied across adult populations and various health conditions, including mental health. It captures five areas of daily life, namely, mobility, self-care, usual activities, pain/discomfort, and anxiety/depression. Each response is rated on five ordered levels of functioning.
- The Recovering Quality of Life-10 (ReQoL-10)^44,45^ will be used to evaluate mental health specific HRQoL. The ReQoL-10 is a preference-based instrument designed for individuals, aged 16 years and above, who live with diverse mental health challenges, including anxiety and depression. It assesses multiple facets of recovery, such as activity, hope, belonging, self-perception, well-being, autonomy, and physical health.
- Client Service Receipt Inventory (CSRI)^39^. An adapted version of the CSRI will be employed to capture health and social care resources used by participations (e.g. utilisation of primary and secondary care services, medication use), as well as time and cost associated to travel to treatment-related appointments. This measure has been shown to align well with case records^46^.
- Institute of Medical Technology Assessment (i-MTA) Productivity Cost Questionnaire (iPCQ)^47^. A modified version of the iPCQ will be used to collect information about participants’ time off from paid and unpaid work (e.g. voluntary activities) due to their mental health problems.
- “Ad-hoc” form/logs. These tailored questionnaires will collect information on resources required to set up and support the delivery of the outdoor swimming courses (e.g. insurance and safety equipment). They will also contain bespoke logs for swimming coaches and their supervisors to record the time they devote to training, preparing, and delivering the outdoor swimming sessions.

#### Intervention evaluation measures and tools

Session attendance will be recorded as the number of outdoor swimming sessions attended by each participant (range: 0–8). Treatment completion will be defined as attendance at a minimum of 50% of sessions, equivalent to at least four sessions. The number and causality of any serious adverse events (SAEs) and adverse events will be systematically documented and reported throughout the trial.

#### Qualitative data collection tools

Qualitative data collections tools will be developed iteratively throughout the project. All participants in the intervention arm will be invited through weekly emails during the swim intervention to complete diaries about their swim experiences. The researchers undertaking the analysis of the diaries will not know the identity of the participants who completed them. Swim along interviews, recorded using a waterproof camera (Go-Pro), will be undertaken at all sites. Further one-to-one interviews will be undertaken in person or using video conferencing after T1. Interviewees will be selected purposively, and participation will be voluntary. Interviewees will be invited to share their diaries with the interviewer, although this will not be a requirement.

### Participant timeline {18}

Table 1 provides the participant timeline for the study.

**Table 1.** Participant timeline: Schedule of enrollment, interventions, and assessments.

| Procedures | Visits |  |  |  |  |  |
| --- | --- | --- | --- | --- | --- | --- |
|  | Screening | Baseline | Allocation | Intervention | Follow up (weeks) |  |
|  |  |  | 0 weeks | 4-12 weeks | 12 | 38 |
| Informed consent | X |  |  |  |  |  |
| Participants details | X |  |  |  |  |  |
| Medical history | X |  |  |  |  |  |
| Physical assessments | X if needed |  |  |  |  |  |
| Demographics |  | X |  |  |  |  |
| Randomisation |  |  | X |  |  |  |
| Intervention:<br>Swim Course |  |  |  | X |  |  |
| Control: Usual care | X |  |  |  |  | X |
| **Intervention arm Assessments:** |  |  |  |  |  |  |
| Swim along interviews (selected sites only) |  |  |  | X (weekly) |  |  |
| Swim Coach Survey |  |  |  | X (weekly) |  |  |
| Participant Diaries |  |  |  | X (weekly) |  |  |
| Swimming Course Questionnaire (intervention arm only) |  |  |  |  | X |  |
| **Study Assessments:** |  |  |  |  |  |  |
| PHQ-9 | X | X |  |  | X | X |
| Swimming Experience Questionnaire |  | X |  |  | X | X |
| GAD 7 |  | X |  |  | X | X |
| EQ5D |  | X |  |  | X | X |
| ReQoL |  | X |  |  | X | X |
| FFMQ-15 |  | X |  |  | X | X |
| CSRI |  | X |  |  | X | X |
| iPCQ |  | X |  |  | X | X |
| Your Health and Your Work |  | X |  |  | X | X |
| Use of Services Questionnaire (parts 1,2,3) |  | X |  |  | X |  |
| Social Prescribing questionnaire |  | X |  |  | X |  |

### Sample size {19}

A cohort study of 400 primary care patients^48^ found a threshold score of 1.7 on the PHQ-9 between ‘feeling better’ and ‘feeling the same’. This is our Minimal Clinically Important Difference (MCID). The pooled baseline SD for PHQ-9 in OUTSIDE 1^34,35^ was 5.23. Using these estimates, we determined an effect size of 1.7/5.23 = 0.325 for OUTSIDE 2.

OUTSIDE 2’s required sample size was estimated using Stata’s *clsampsi* routine^49^ with the following parameters: intracluster correlation coefficient (ICC) of 0.01^50^, clustering in the swim intervention arm only, cluster size of 8 swimmers (however this will vary for pragmatic reasons between a swim intervention group size of between 5 and 11, aiming for an average of around 8), 90% power, two-sided test with 5% alpha, and 1:1 allocation. This yielded n=216 per arm, N=432 total.

For repeated measures, a multiplying factor (R) was applied using Equation 6.7 in Machin et al^51^ with 1 pre-randomisation, 1 post-randomisation observation, and a correlation of 0.52 for baseline and week-8 follow-up PHQ-9 scores^35^. This yielded R=0.7296, reducing the sample size to n=316. Accounting for a 25% attrition rate^35^, the required sample size is n=211 per arm, N=422 total.

However, anticipating cancellation of 3-4 swim courses due to last minute cancellations, unavoidable or unpredictable factors (e.g. poor weather or poor water quality meaning swim coaches are unable to conduct sessions) we will recruit to allow for this in addition to allowing for study attrition. This results in n=240 in the swim intervention (30 courses of 8 people on average) and control arms, or N=480.

### Recruitment {20}

Based on the OUTSIDE 1 feasibility RCT^34,35^, participants for OUTSIDE 2 will be recruited through the following mechanisms:

1. Develop promotional material that includes people under-represented in outdoor swimming in particular people of colour, and men and advertise in groups that represent these communities.
2. Advertise on social media, including Facebook and Instagram and local online forums, with links to the OUTSIDE 2 trial website, films and podcasts.
3. Display posters and flyers in swim venues, local GPs, social prescribing and talking therapy services.
4. Identify and liaise with social prescribers where possible and ask them to invite potential participants to self-refer and emphasise our need to recruit a diverse participant sample (e.g. males, ethnic minorities, people with a disability, people with lower educational attainment, mix of socio-economic backgrounds, neurodiverse).
5. Those registered with the National Institute of Health and Care Research’s (NIHR) ‘Be Part of Research’ service who meet the OUTSIDE 2 study eligibility criteria.
6. Those registered with the GLAD study team (https://gladstudy.org.uk/about/) who requested to be informed of further research and also meet the OUTSIDE 2 study criteria.
7. Database search and mail out from GP practices close to swim sites that are part of NIHR’s Research Delivery Network. The searches will yield potential participants, participating practices will be asked to send an email or text, providing information about the trial, to all eligible patients.
8. In underserved communities, local trusted community connectors will be sought to help advertise the trial. e.g. in among different ethnic minority communities and Lesbian, Gay, Bisexual, Transgender, Queer, or Questioning and others (LGBTQ+) communities. We will use the demographic data following the first phase of recruitment to inform the selection of two bespoke courses for underserved groups with specific protected characteristics. These swim courses would be solely for participants from these communities to encourage participation.

#### Recruitment process

Participants will be directed first to the study website (<u>outside2.co.uk</u>), which will act as the main source of information about the trial. Interested individuals will be able to contact the study team via an embedded expression of interest form. Following this, the participant information sheet and consent form will be emailed to prospective participants, and an eligibility appointment will be scheduled. The consent process will adhere to National Institute for Health Research (NIHR) Good Clinical Practice (GCP) guidelines, and all research team members responsible for obtaining consent will have completed GCP training.

Eligibility assessments will be conducted by research assistants (RAs) via telephone or videoconference, no sooner than 24 hours after the participant information sheet has been sent. During this appointment, participants will have the opportunity to ask questions about the study and will be asked to provide verbal consent to proceed with eligibility screening. The RA will assess inclusion and exclusion criteria, and the study medical team will be consulted in cases where health history, suicide risk, or current psychosis may indicate exclusion.

If eligibility is confirmed, participants will be sent the baseline survey (T0), which will be administered no more than four weeks prior to the first swimming session. The survey may be completed independently online or with support from an RA via videoconference or telephone, if requested.

Upon completion of the baseline survey, participants will be enrolled in the study and randomised to either the outdoor swimming intervention arm or the usual care control arm. Participants will be notified of their allocation, and a letter will be sent to their general practitioner (GP) to inform them of the participant’s inclusion in the trial.

### Randomisation

#### Sequence generation {21a, 21b} and Concealment mechanism {22}

The Brighton and Sussex Clinical Trials Unit (BSCTU) will set up the online randomisation process using the online randomisation module of REDCap™ with stratification by site, using permuted blocks of randomly varying length. For safety reasons, maximum capacity at each site is determined by staff to swimmer ratios. The allocation mechanism makes it impossible to guess the next allocation therefore if the intervention group is at capacity, the site will be closed to further randomisations.

#### Implementation {23}

The trial manager (TM) at the BSCTU will communicate the randomisation outcome to the designated unblinded research assistant (RA). Participants will then be notified of their allocation via email. The unblinded RA will subsequently contact the assigned swim coach, providing relevant participant details including contact information, emergency contact details, previous swimming experience, and any additional information the participant has requested to be shared. The swim coach will then initiate contact with the participant to introduce themselves and provide details about the introductory session and the scheduled swim sessions.

### Assignment of interventions: Blinding

#### Who will be blinded and how it will be achieved {24a, 24b}

Two RAs will remain blinded until the final swim course starts, after that point one RA will remain blinded. The Trial Statistician, CTU Statistician and Health Economists will be blind to participant allocation until the final data are collected and entered. The Statistical Analysis Plan (SAP) has been signed off ahead of the final data collection (see Appendix 2 for SAP) and a Health Economics Analysis Plan (HEAP)^52^ will be signed off before the final data are collected. Blinding will be maintained through several measures: (1) as in OUTSIDE 1, the majority of participants will complete surveys online without a blinded research assistant (RA) present; (2) participants will be reminded at the start of each assessment interview not to reveal their allocation arm; (3) blinded research team members will be kept separate from any discussions in which participant allocation could be inferred; (4) researcher access to electronic health records will be limited; and (5) office arrangements and administrative procedures will be organised to separate blinded and unblinded staff. Awareness and education around maintaining blinding will be promoted throughout the study. To assess the effectiveness of blinding, the RA who conducted each participant’s eligibility assessment will be asked to guess the participant’s allocation arm after the T2 assessment. Any reported breaches of blinding will be recorded.

#### Procedure for unblinding if needed {24c}

Unblinding will follow the trial team’s operating procedures. This policy outlines which team members will remain blinded, the circumstances under which unblinding may occur, and the processes for managing unblinding events. These include training requirements, controlled access to electronically stored unblinded data, and use of a designated trial email address for handling unblinded queries.If a blinded RA becomes unblinded during an assessment appointment, responsibility for completing the assessment will be transferred to an alternative RA who has no knowledge of the treatment allocation to maintain blinding integrity.

### Data collection and management

No data are associated with this article.

#### Plans for assessment and collection of outcomes {25a}

See Table 1. for the details of data collection at each time point.

##### Screening measures

See above for further details of screening measures.

##### Quantitative data collection

Validated survey information for clinical and health economic outcomes will be collected online at baseline (T0), 12 weeks post randomisation (T1) and at 38 weeks post randomisation (T2) using REDCap™. Survey completion will be prompted by automated e-mail and reminders sent to participants at weekly intervals for up to four weeks and data entry validation will be used to ensure data is entered to minimize missing data. Access to the survey data is limited to blinded members of the team during the data collection and analysis.

##### Qualitative data collection

Diaries will be collected using Qualtrics, with a link sent to each intervention participant shortly after each swim session. Encrypted SD cards will be used for the recording of swim-along interviews. In person and video conferencing interviews will be recorded using video conferencing software.

#### Plans to promote participant retention and complete follow-up {25b}

Efforts will be made to maximise participant engagement with follow-up assessments through the use of reminder emails and flexible scheduling. Research assistants (RAs) will offer appointments at times that best suit participants and will provide reasonable adjustments, including shorter or split assessment sessions where needed. To support retention, participants will be offered a £20 voucher for each completed survey, with a total of £60 available for completing all three surveys. Retention rates will be reviewed monthly by the trial manager and research team throughout the study. The LEAP will be consulted regularly to offer feedback and recommendations on retention strategies as part of their oversight role.

In line with recommendations from PPI representatives, participants randomised to the control arm will be offered the opportunity to take part in an introductory outdoor swimming course upon completion of the trial. This approach is intended to enhance acceptability of randomisation and reduce attrition, consistent with established practices for improving retention^53^.

#### Data management {26}

The research team will comply with the standards and principles set out in the Sussex Partnership NHS Foundation Trust Policy for Data Protection, Security, and Confidentiality. This policy aligns with current legislative and regulatory requirements, including the Caldicott Report (1997), the British Standard for Information Security (ISO/IEC 27002), the General Data Protection Regulation (GDPR, 2018), and the Sussex Partnership NHS Foundation Trust Research Policy (2021). Data management and analysis will follow the CTU’s Standard Operating Procedures (SOPs), and data quality assurance will be overseen through the CTU’s Data Management Plan, which can be requested from the corresponding author.

Participants will be fully informed, via the study consent form, of the conditions under which their anonymised data may be reused or shared for future research purposes. No identifiable information will be shared with individuals outside of the research team.

All research activities will adhere to these standards and will be reviewed and approved by an NHS Research Ethics Committee, the Health Research Authority, and relevant Research and Development departments in accordance with the UK Framework for Health and Social Care Research (2017). All members of the research team and collaborators across participating NHS Trusts and universities involved in data collection, processing, or sharing will have completed Information Governance training. Data management will be a standing item at monthly research team meetings to ensure continuous oversight and compliance.

#### Confidentiality {33}

Consent forms will be stored in password-protected files on secure NHS Trust servers. During the eligibility assessment, participants’ names will be recorded on the consent forms. Following this, each participant will be assigned a unique study identification code by the RA. Identifiable information, including names and study IDs, will be stored separately. A secure participant identification log linking personal information to study codes will be stored in a password-protected file accessible only to authorised members of the research team.

REDCap will be used to collect all quantitative data which will be pseudonymised using study identification codes. These data will be securely stored on AROTECH data centre servers, with restricted access to essential research members only and routine back up procedures. Attendance data will be collected weekly from swim coaches using structured forms, which will include session activities and be stored on NHS Trust servers in password-protected files.

Diary entries will be collected via Qualtrics and downloaded to NHS Trust servers for secure storage and analysis. Swim-along video interviews will be recorded onto encrypted SD cards, while post-course interviews will be conducted via secure video conferencing software. All recordings will be uploaded to password-protected NHS Trust servers and stored in encrypted files. Once uploaded, recordings will be deleted from the original devices and SD cards will be reformatted. Interview transcripts will be stored in password-protected files and will be anonymised, with no identifiable information included.

All electronically stored data, including questionnaire scores, video files, and transcripts will be anonymised and maintained in encrypted formats on secure shared drives. Access to these drives will require a username and password, and will be restricted to members of the research team. Personal information will be securely destroyed once it is no longer required for the study.

## Statistical methods

### Statistical Analysis Plan (SAP) {5}

A detailed SAP (see Appendix 2) has been written by the CTU Statistician and Trial Statistician, agreed with the research team and will be reviewed by the Trial Steering Committee (TSC).

### Statistical methods for primary and secondary outcomes {27a, 27b}

Participant flow through the trial will be presented in accordance with the CONSORT-SPI 2018 extension for reporting social and psychological intervention trials^54,55^. Baseline characteristics and outcome measures will be summarised by trial arm using descriptive statistics appropriate to the distribution of each variable. The primary analysis will adhere to intention-to-Treat (ITT) principles where all participants included in the analysis will be as per their random allocation. The primary outcome will be analysed using a linear mixed effects model with: treatment arm (swimming + usual care or usual care alone), time (T1 or T2), and treatment arm × time interaction added as fixed factors; baseline PHQ-9 and GAD-7 scores and reports of at least one previous episode of depression (derived from the MINI Depression Module) will be covariates; site/swim-group will be added as a random factor (random intercept) and random subject effects will be included to account for correlation between repeated measures on individual participants with a small sample adjustment. The site/swim group factor categories indicate the site and swim group which will simultaneously account for year. The primary endpoint, the adjusted between-arm difference in mean PHQ-9 at T1, will be estimated with corresponding 95% confidence interval and p-value. All p-values will be significant at the 5% alpha level using two-sided tests. PHQ-9 at T2 and secondary outcomes at T1/T2 will be analysed using similar methods. Standardised (Cohen’s d) between-group treatment effect sizes will be calculated for each outcome at T1 and T2. A safety and lasting negative effects summary will be conducted for all participants by trial arm. Where swim groups are cancelled due to last minute or unpredictable problems (poor water quality or weather which means the swim coaches cannot conduct the course) the removal of these data would form part of a sensitivity analysis. Stata 19 will be used for all analyses.

#### Alternative analyses

If the model for the primary analysis does not converge then we will re-run the primary analysis model with site included as a fixed effect.

### Interim analyses {28b}

There are no planned interim analyses.

### Methods for additional analyses (e.g. subgroup analyses) {27d}

#### Subgroup analyses

Subgroup analyses will explore whether estimated treatment effects vary by, gender, ethnicity, and type of swim location (e.g. lido, sea, lake). The size of the groups, effect size, 95% CI and p-values will be stated. This trial is not powered to detect subgroup effects and so caution will be taken when interpreting the results.

#### Health Economic Analysis

The trial economic components will be detailed in a Health Economics Analysis Plan (HEAP)^52^, which will be completed prior to any data analysis. The primary economic evaluation will be a cost-utility analysis (CUA), with a cost-effectiveness analysis (CEA) conducted as secondary economic evaluation. Base-case analyses for both will be undertaken from the NHS and Personal Social Services (PSS) perspective, in line with the National Institute for Health and Care Excellence (NICE) guidelines^56^, and will follow an intention-to-treat (ITT) approach.

Sensitivity analyses will adopt a wider societal perspective to capture the broader economic impact of mental health problems, such as productivity losses. The economic components will follow established best-practice guidelines for performing economic evaluations and reporting their results^56,57^. Costs and outcomes will be compared across trial arms at the 38-week follow-up. Missing data will be addressed with multiple imputation methods^58,59^, with primary and secondary economic analyses relying on imputed data. In the CUA, health outcomes will be expressed as QALYs gained, as estimated from the EQ-5D-5L^43,60^ and the ReQOL^44,45^ instruments. In the CEA, effectiveness will be assessed using the PHQ-9 (primary clinical outcome).

Resource use data will be collected for each participant using an adapted version of the CSRI questionnaire^39^ and will include treatment as usual and medications, resources linked to the outdoor swimming intervention (for the intervention arm), other health and social care services, and broader societal resources. These data will be obtained from the participants, with further data related to attendance and other swimming costs gathered from bespoke logs completed by swimming coaches and their supervisors. Total cost per participant will be calculated by applying units costs of resources to their recorded quantities and then summing across cost categories. Unit costs will be mainly sourced from established local and nationals collections (e.g. PSSRU; National Cost collection for the NHS^61^, Prescription Cost Analysis (PCA)^62,63^. Costs will be reported in pounds sterling at current prices and adjusted for inflation if necessary. Given the short duration of the trial, neither costs nor outcomes will be discounted. The economic evaluation findings will be presented in the CUA as incremental cost per QALY gained, and in the CEA as incremental cost per variation in the PHQ-9 outcome. Uncertainty will be quantified and illustrated with cost-effectiveness acceptability curves^64^. Sensitivity analyses will assess robustness and generalisability of economic results. They will include, for instance, adopting a societal perspective; complete case analysis; per-protocol analysis. Explorative subgroup analyses may be also performed in line with statistical analyses.

#### Qualitative analysis of participants’ experience

Interviews will be transcribed and, together with diaries, analysed using reflexive thematic analysis^65^. Data will be systematically coded and categorised to develop themes that reflect and illuminate patterns with the data. NVivo will be used to aid this process. Reflexivity, the process of critically reflecting on your position as a researcher and on the research process, will be supported by keeping reflexive journals, regular discussions within the qualitative research team, and sharing, and inviting feedback, from the wider team and members of the LEAP as the analyses develop. Due to the large data capture the iterative analysis is likely to move to a deductive analysis that will enable further consolidation of the themes identified.

### Methods in analysis to handle protocol non-adherence and any statistical methods to handle missing data {27c}

As a sensitivity analysis, a secondary complier average causal effect (CACE)^66^ analysis will be conducted to assess the impact of compliance (attending 4 or more sessions out of 8) on the primary outcome. Using a Structural Equation Model (SEM)^67^, a two-stage instrumental variables regression approach will be used with treatment assignment as the instrumental variable and adjustments for all covariates and fixed factors included in the primary analysis; robust standard errors will be used to account for within-site clustering. Compliance will be defined as i) a continuous measure of the number of swim sessions attended and then ii) a dichotomous measure indicating that four or more swim sessions were attended.

Missing item-level data will be addressed following the outcome scale’s guidance, or as appropriate, using appropriate statistical techniques for handling missing data. Linear mixed models will use maximum likelihood estimation, which is effective for handling missing data under the assumption that data is Missing At Random.

### Oversight and monitoring

#### Composition of the coordinating centre and trial steering committee {3d}

A Trial Steering Committee (TSC) will be established in accordance with Medical Research Council (MRC) guidelines^69^ and will be chaired by a senior clinical academic. At least two independent experts and two independent lay members will sit in the TSC and will meet at least once a year to provide oversight of the trial, review adherence to the protocol, and offer independent guidance on trial conduct.

A LEAP will provide PPI oversight. All LEAP members will receive appropriate training to support meaningful involvement and will meet throughout the study. Their consultation will focus on key areas including development of accessible study materials including informed consent documentation, provide suggestions for recruitment and retention strategies, interpretation of findings, and dissemination planning. An involvement log will be maintained to document the influence of LEAP contributions on trial decisions and processes.

#### Composition of the data monitoring committee, its role and reporting structure {28a}

The Data Monitoring and Ethics Committee (DMEC) will report directly to the TSC and will meet at least once a year. The DMEC will have unrestricted access to unblinded trial data and will receive reports on adverse events. DMEC members will be independent of both the TSC and trial applicants. The DMEC will be chaired by an independent senior statistician and will include an additional independent clinician and senior academic. The Chief Investigators (CIs), Trial Manager (TM), and trial statistician will attend portions of the DMEC meetings to present reports but will not serve as members of the committee.

#### Adverse event reporting and harms {17}

Any deterioration in a participant’s mental or physical health requiring an unexpected contact with a healthcare professional (unless this constitutes an SAE, see below) will be recorded as an adverse event (AE), regardless of whether it is judged to be related to the study intervention.

Serious Adverse Events (SAEs) are defined as events that are life-threatening, resulting in death, require inpatient hospitalisation, a prolongation of existing hospitalisation, or lead to significant or persistent incapacity/disability, congenital abnormality or a birth defect.

The number of events (both by event and by individuals) and nature of all AEs and SAEs reported to blinded and unblinded members of the research team will be documented and summarised.

The reporting period for SAEs and AEs will span from giving informed consent to the last follow-up assessment at 38 weeks post-randomisation. Each adverse event will be systematically recorded and reviewed by a Co-CI. In cases where an AE is classified as a serious adverse event (SAE), it will be assessed for causality and expectedness by one of the Chief Investigators in consultation with an independent clinical expert. Details of all SAE reviews (including their number, nature, and outcomes) will be communicated to the TSC, which will advise on any consequent actions. Serious adverse events will also be reported to the Sponsor, the Data Monitoring and Ethics Committee, and the NHS Research Ethics Committee, where appropriate.

Significant risk to self (e.g. risk of suicide) or others that is identified at any point from enrolment to T2 will be discussed immediately with mental healthcare professionals in the study team and a proportionate response taken in line with good clinical practice. This may include, depending on the nature and immediacy of risk, alerting the person’s GP the same day, arranging for the person to be accompanied to their nearest Accident and Emergency Department or speaking to the person on the phone whilst arranging for an ambulance to attend.

#### Frequency and procedures for monitoring trial conduct {29}

The TSC and DMEC will operate independently of the trial investigators and sponsor to ensure the study is conducted to the highest standards. Oversight will include monitoring trial conduct, adherence to the protocol, and participant safety. Audit, monitoring, and inspection activities will be carried out in accordance with the procedures of the trial sponsor, Sussex Partnership NHS Foundation Trust (SPFT), and the CTU.

### Ethics and Dissemination plans

#### Ethics approval and consent to participate {30}

The study is sponsored by Sussex Partnership NHS Foundation Trust. NHS Research Ethics Committee (London - Hampstead, reference number 23/LO/0942), Health Research Authority and local Research Governance approval were granted before the commencement of the trial (IRAS 328871 09/01/2024 Protocol v1.3, at time of writing v11 08/07/2025). Participants will provide written informed consent prior to the completion of any study procedures.

#### Plans for communicating important protocol modifications to relevant parties {31}

Protocol amendments will be formally submitted for review and approval by the Research Ethics Committee and the Health Research Authority. Once approved, these changes will be incorporated into the updated study protocol and recorded in the ISRCTN trial registration.

#### Dissemination {8}

Trial findings will be disseminated through peer-reviewed scientific publications, regardless of outcome, including clinical and cost effectiveness, and insights from the qualitative analysis of participants’ experiences. Findings will also be shared with participants and relevant patient organisations. Members of the LEAP will actively contribute to dissemination activities, including the use of social media and preparation of a lay summary for non-academic publication. Trial results will be presented at public engagement events, as well as at local, national, and international academic and professional conferences. Additionally, findings will also be published on the sponsor’s website to ensure broad accessibility.

## Discussion

There is a growing body of evidence indicating potential positive effects of outdoor swimming for individuals experiencing depression. To ensure a thorough assessment of the potential clinical and cost effectiveness and safety of outdoor swimming courses, rigorous mixed methods randomised controlled trials are necessary. This protocol is for a full-scale RCT of an 8-session outdoor swimming course for adults with depression with an embedded economic evaluation to assess value for money and a qualitative study exploring participants experiences developed from the OUTSIDE 1 feasibility RCT^37^ that indicated a definitive RCT was feasible and warranted^35^.

This study will determine if the 8-week outdoor swimming course conducted by experienced and trained outdoor swimming coaches is safe and if it is clinically and cost effective for adults experiencing depression. If findings demonstrate clinical effectiveness and safety in comparison to usual care, we will work with policy makers and providers to support wide-scale roll out of the course so that people living with the devasting effects of depression have the opportunity to benefit from an evidence-based intervention. If findings demonstrate cost-effectiveness, this will further support national roll out by showing that the intervention is good value for money and potentially cost saving to the wider healthcare sector. Furthermore this may pave the way for other outdoor activities to support people with poor mental health to be developed and evaluated as interventions.

### Trial status

Recruitment to the trial commenced in March 2024. Recruitment is planned to continue until June 2025 and follow-up data collection until February 2026.

## Supporting information

SPIRIT Checklist

Statistical Analysis Plan

## Data Availability

No underlying data or extended are associated with this article. The SPIRIT (2025) checklist for OUTSIDE 2: Outdoor swimming as a nature-based intervention for depression is available at https://doi.org/10.25377/sussex.3144518868

## Abbreviations

AE: Adverse Event
BSCTU: Brighton and Sussex Clinical Trials Unit
CI: Chief Investigator
CSRI: Client Service Receipt Inventory
DMEC: Data Monitoring and Ethics Committee
EQ-5D-5L: Euroqol 5-Dimension 5-Level instrument
FFMQ-15: Five-Facet Mindfulness Questionnaire
GAD-7: General Anxiety Disorder-7
GCP: Good Clinical Practice GP General Practitioner
HRQoL: Health-related quality of life
ISRCTN: International Standard Randomised Controlled Trial Number
LEAP: Lived Experience Advisory Group
MINI: Mini International Neuropsychiatric Interview
NICE: National Institute for Health and Care Excellence
PHQ-9: Patient Health Questionnaire-9
PIC: Participant Identification Centre
PIS: Participant Information Sheet
PPI: Patient and Public Involvement
RCT: Randomised Control Trial
REC: Research Ethics Committee
ReQoL-10: Recovering Quality of life - 10 items
SAE: Serious Adverse Event
SMG: Study Management Group
SOP: Standard Operating Procedure
SPFT: Sussex Partnership Foundation Trust
TSC: Trial Steering Committee

## Declarations

### Data availability {6}

An anonymised dataset will be deposited in the University of Sussex research repository to support open access and facilitate secondary analysis by other researchers. Participants will be informed of the potential for anonymised data reuse and sharing as part of the consent process. No identifiable information will be shared outside the research team, and all data handling will comply with relevant data protection and confidentiality standards.

### Underlying and Extended data

No underlying data or extended are associated with this article.

### Reporting Guidelines

Figshare. SPIRIT (2025) checklist for OUTSIDE 2: Outdoor swimming as a nature-based intervention for depression, https://doi.org/10.25377/sussex.31445188^68^.

This project contains the following underlying data:

- SPIRIT (2025) checklist for OUTSIDE 2: Outdoor swimming as a nature-based intervention for depression protocol

Data are available under the terms of the <u>Creative Commons Zero “No rights reserved” data waiver</u> (CC0 1.0 Public domain dedication).⍰

## Acknowledgements

We wish to acknowledge the significant contribution to the study made by the swim coaches and their support teams (Vicki McCreadie, Gilly McArthur, Jonathan Cowie, Lauren Munro-Bennett [Rayrigg Meadow, Lake Windermere]; Louise Parker and Jacklyn McCourt [Roker Beach, Sunderland]; Christine Bradley and Andrea Turner [Ilkley Lido, Ilkley] Jo Rowbotham, Colin McCard, Fiona Hanik [West Kirby Marine Lake, Wirral]; Karen and David Quartermain [USwim, Salford Quays]; Justine Wales and Cat Wynn [Whole Health Swim Village, Nottingham]; Gem Lamsdale, Emily Lang, Sophy Howard and Penny Wilson [Watersedge, Bishampton, Worcester]; Jonathon Hardy, Colin Campbell and Sophie Etheridge [Jesus Green Lido, Cambridge]; Paul Slade and Mo Silcock [Chalkwell Beach, Leigh-on-Sea]; Maggy Blagrove, Liz Dunn and Melissa Oghre [Open Minds Active CIC, West Country Water Park, Bristol]; Carine Evans and Janet Moore [Taplow Lake, Maidenhead]; Paul MacKenzie and Jude Palmer [Embrace Adventures CIC, Quay Lake, Camberley]; Nina Yates [Lymington Sea Baths, Lymington]; Claire Anderson and Omie Dale [Parliament Hill Lido and West Reservoir, London]; Hazel Fulker and Steve [Loddington Farm Lake, Maidstone]; Fiona Mildner and Marianne Clarke [Sea Sure, Sea Lanes, Brighton]; Lawrence Naested, Kath Lambert [Widewater Beach, Shoreham-by-Sea]; Jo Cox, Will Verling, Vivienne Bryans, Nicky Painter and Simon Lancaster [CHILL CIC, Avon Beach, Christchurch]; Lucy Green and Lorraine Brown [Beyond the Wave CIC, Penzance and Falmouth]), our Research and Development team (Matt Smith, Yvette Wagner, Lucy Brocklesby, Ian Lasky, Craig Peachy and Jane Shiers), research assistants and clinical research coordinators (Lily Holland, Diana Phillips, Ruby Warden, Laura Hernshaw, Emily Budden, Hannah Scannell, Rhianon Potter and Adam Jones). We also appreciate the advice and assistance offered by the Trial Steering Committee, the DMEC and the experts by experience on the LEAP and Chloe Elsby-Pearson and Lucy Walsh who facilitated the sessions.

We thank the NIHR BioResource volunteers for their participation, and gratefully acknowledge NIHR BioResource centres, NHS Trusts and staff for their contribution. We thank the National Institute for Health and Care Research, NHS Blood and Transplant, and Health Data Research UK as part of the Digital Innovation Hub Programme. The views expressed are those of the author(s) and not necessarily those of the NHS, the NIHR or the Department of Health and Social Care. We also thank the NIHR Be Part of Research team for their support promoting the study to potential participants local to the study locations. The views expressed are those of the author(s) and not necessarily those of the NHS, the NIHR or the Department of Health and Social Care.

## References

1. Lim GY, Tam WW, Lu Y, Ho CS, Zhang MW, Ho RC. Prevalence of depression in the community from 30 countries between 1994 and 2014. Sci Rep. 2018 Feb 12;8(1):2861. doi:10.1038/s41598-018-21243-x

2. Office for National Statistics. Cost of Living and Depression in Adults [Internet]. Great Britain: ONS; 2022 [updated 2022 Dec 06; accessed 2025 May 18]. Available from: https://www.ons.gov.uk/peoplepopulationandcommunity/healthandsocialcare/mentalhealth/articles/costoflivinganddepressioninadultsgreatbritain/29septemberto23october2022

3. NHS Business Services Authority. NHS releases 2023/24 mental health medicines statistics for England [Internet]. Newcastle: NHS; 2024 [updated 2024 Jul 11; accessed 2025 May 18]. Available from: https://media.nhsbsa.nhs.uk/press-releases/5171d616-95ea-4282-959b-15f8bfed6a0f/nhs-releases-2023-24-mental-health-medicines-statistics-for-england

4. American Psychological Association. Clinical Practice Guideline for the Treatment of Depression Across Three Age Cohorts [Internet]. Washington, DC: APA; 2019 [updated 2019 Feb; accessed 2025 May 13]. Available from: https://www.apa.org/depression-guideline/guideline.pdf

5. National Institute for Health and Care Excellence. Depression in Adults: Treatment and Management [Internet]. London: NICE; 2022. [updated 2022 Jun 29; accessed 2025 May 18] Available from: https://www.nice.org.uk/guidance/ng222

6. Cipriani A, Furukawa TA, Salanti G, Chaimani A, Atkinson L, Ogawa Y. Comparative efficacy and acceptability of 21 antidepressant drugs for the acute treatment of adults with major depressive disorder: a systematic review and network meta-analysis. Lancet. 2018 Apr 7;391(10128):1357–66. doi:10.1016/S0140-6736(17)32802-7

7. Gartlehner G, Wagner G, Matyas N, Titscher V, Greimel J, Lux L, Gaynes BN, Viswanathan M, Patel S, Lohr KN. Pharmacological and non-pharmacological treatments for major depressive disorder: review of systematic reviews. BMJ Open. 2017 Apr 27;7(6):e014912. doi:10.1136/bmjopen-2016-014912

8. Gartlehner G, Thieda P, Hansen RA, Gaynes BN, Deveaugh-Geiss A, Krebs EE, Lohr KN. Comparative risk for harms of second-generation antidepressants: a systematic review and meta-analysis. Drug Saf. 2008;31(10):851–65. doi:10.2165/00002018-200831100-00004

9. NHS Digital. NHS Talking Therapies Monthly Statistics Including Employment Advisors, Performance December 2024 and Quarter 3 2024/25 Data [Internet]. London: NHS Digital; 2025. [updated 2025 Feb 13; accessed 2025 May 20] Available from: https://digital.nhs.uk/data-and-information/publications/statistical/nhs-talking-therapies-monthly-statistics-including-employment-advisors/performance-december-2024-and-quarter-3-2024-25-data

10. World Health Organization. Guidelines on Physical Activity and Sedentary Behaviour [Internet]. Geneva: WHO; 2020. [updated 2020 Nov 25; accessed 2025 Oct 20]. Available from: https://www.who.int/publications/i/item/9789240015128

11. Tavares VD de O, Williams JVA, Sharifi V, Bulloch A, Dimitropoulos G, Galvão-Coelho NL, Patten S. Physical activity and mortality in the general population with and without major depression. Acad Ment Health Well-Being. 2024 Sep 15;1(2). doi:10.20935/MHealthWellB7335

12. Cooney GM, Dwan K, Greig CA, Lawlor DA, Rimer J, Waugh FR, McMurdo M, Mead GE. Exercise for depression. Cochrane Database Syst Rev. 2013 Sep 12;2013(9). doi:10.1002/14651858.CD004366.pub6

13. Babyak M, Blumenthal JA, Herman S, Khatri P, Doraiswamy M, Moore K, Craighead WE, Baldewicz TT, Krishnan KR. Exercise treatment for major depression: maintenance of therapeutic benefit at 10 months. Psychosom Med. 2000 Seo-Oct;62(5):633–8. doi:10.1097/00006842-200009000-00006

14. NHS England. Health Survey for England 2021 Part 2 [Internet] London: NHS England; 2023. [Updated 2023 May 16, accessed 2025 May 20]. Available from: https://digital.nhs.uk/data-and-information/publications/statistical/health-survey-for-england/2021-part-2/physical-activity

15. Geary RS, Thompson D, Mizen A, Akbari A, Garrett JK, Rowney FM, et al. Ambient greenness, access to local green spaces, and subsequent mental health: a 10-year longitudinal dynamic panel study of 2.3 million adults in Wales. Lancet Planet Health. 2023 Oct;7(10):e809–18. doi:10.1016/S2542-5196(23)00212-7

16. Geary R, Thompson D, Garrett J, et al. Green–blue space exposure changes and impact on individual-level well-being and mental health: a population-wide dynamic longitudinal panel study. Public Health Res. 2023 Oct;11(10). doi: 10.3310/LQPT9410

17. Roba HS, Kolbe-Alexander T, Baliunas D, Biddle SJH. Associations between green space, blue space and mental health outcomes in regional Australia: cross-sectional and longitudinal analyses. J Affect Disord. 2025 Jun 16;389:119685. doi:10.1016/j.jad.2025.119685

18. Sport England. Active Lives Adult Survey (November 2023–24) Report [Internet]. Loughborough: Sport England; 2025. [updated 2025 Apr, accessed 2025 May 18] Available from: https://sportengland-production-files.s3.eu-west-2.amazonaws.com/s3fs-public/2025-04/ActiveLivesAdult-Nov23-24_V9-23-04-25-10-03-03-02.pdf https://sportengland-production-files.s3.eu-west-2.amazonaws.com/s3fs-public/2025-04/ActiveLivesAdult-Nov23-24_V9-23-04-25-10-03-03-02.pdf

19. Denton H, Aranda K. The wellbeing benefits of sea swimming: is it time to revisit the sea cure? Qual Res Sport Exerc Health. 2019 Sep 2;12(5):647–63. doi:10.1080/2159676X.2019.1649714

20. Massey H, Kandala N, Davis C, Harper M, Gorczynski P, Denton H. Mood and well-being of novice open water swimmers and controls during an introductory outdoor swimming programme: a feasibility study. Lifestyle Med. 2020 Nov 10;1(2):e12. doi:10.1002/lim2.12

21. Burlingham A, Denton H, Massey H, Vides N, Harper CM. Sea swimming as a novel intervention for depression and anxiety: a feasibility study. Ment Health Phys Act. 2022 Oct;23:100472. doi:10.1016/j.mhpa.2022.100472

22. Massey H, Gorczynski P, Harper CM, Sansom L, McEwan K, Yankouskaya A, Denton H. Perceived impact of outdoor swimming on health: web-based survey. Interact J Med Res. 2022 Jan 4;11(1):e25589. doi:10.2196/25589

23. Overbury K, Conroy BW, Marks E. Swimming in nature: a scoping review of mental health and wellbeing benefits. J Environ Psychol. 2023 Aug 5;90:102073. doi:10.1016/j.jenvp.2023.102073

24. Tipton MJ, Collier N, Massey H, Corbett J, Harper M. Cold water immersion: kill or cure? Exp Physiol. 2017 Nov 1;102(11):1335–55. doi: 10.1113/EP086283

25. Tipton M, Massey H, Mayhew A, Morgan P. Cold water therapies: minimising risks. Br J Sports Med. 2022 Dec;56(23):1332–4. doi:10.1136/bjsports-2022-105953

26. Faivre-Rampant V, Hingrand C, Mezanger A, Saloux E, Ollitrault P, Alvado S, Normand H, Mekjavic IB, Collet T, Mauvieux B, Drigny J, Hodzic A. Cardiac electrical and functional activity following an outdoor cold-water swimming event. J Therm Biol. 2024 Oct; 103996. doi:10.1016/j.jtherbio.2024.103996

27. Bell SL, Foley R, Houghton F, Maddrell A, Williams AM. From therapeutic landscapes to healthy spaces: a scoping review. Soc Sci Med. 2018 Jan; 196:123–30. doi:10.1016/j.socscimed.2017.11.035

28. Environment Agency. The Social Benefits of Blue Space: A Systematic Review [Internet]. Bristol: Environment Agency UK; 2020. [Updated 2020 Oct, Accessed 2025 May 21]. Available from: https://assets.publishing.service.gov.uk/media/5f900877d3bf7f5d532ca0a5/Social_benefits_of_blue_space_-_report.pdf

29. Job S, Heals L, Obst S. Oceans of opportunity for universal beach accessibility: an integrated model for health and wellbeing. Aust N Z J Public Health. 2022 Jun; 46(3):252–4. doi:10.1111/1753-6405.13246

30. Thomas F. The role of natural environments within women’s everyday health and wellbeing in Copenhagen, Denmark. Health Place. 2015 Sep;35:187–95. doi: 10.1016/j.healthplace.2014.11.005

31. Hignett A, White M, Pahl S, Jenkin R, Froy M. Evaluation of a surfing programme designed to increase well-being among at-risk young people. J Adventure Educ Outdoor Learn. 2018 May 19;18(1):53–69. doi: 10.1080/14729679.2017.1326829

32. Burdsey D. ‘The foreignness is still quite visible in this town’: multiculture and prejudice at the English seaside. Patterns Prejudice. 2013 Mar 08; 47(2):95–116. doi: 10.1080/0031322X.2013.773134

33. Black Swimming Association. Our Research [Internet]. London: BSA; 2025. [accessed 2025 May 18] Available from: https://thebsa.co.uk/our-research/

34. Massey H, Denton H, Burlingham A, Violato M, Bibby-Jones AM, Cunningham R, Ciccognani S, Robertson S, Strauss C. OUTdoor Swimming as a nature-based Intervention for DEpression (OUTSIDE): study protocol for a feasibility randomised control trial comparing an outdoor swimming intervention to usual care for adults experiencing mild to moderate symptoms of depression. Pilot Feasibility Stud. 2023 Jul 13;9(1):122. doi: 10.1186/s40814-023-01358-3.

35. Massey H, Denton H, Burlingham A, Violato M, Bibby-Jones AM, Cunningham R, Ciccognani S, Robertson S, Jhans A, Pollard J, Yu S, Strauss C. OUTSIDE: OUTdoor Swimming as a nature-based Intervention for DEpression: a feasibility randomised controlled trial. Ment Health Phys Act. 2025; 29:100723. doi: 10.1016/j.mhpa.2025.100723

36. Kroenke K, Spitzer RL, Williams JBW. The PHQ-9: validity of a brief depression severity measure. J Gen Intern Med. 2001 Sep;16(9):606–13. doi:10.1046/j.1525-1497.2001.016009606.x

37. Denton H, Robertson S, Ciccognani S, Meddings S, White P, Elsby-Pearson C, Jhans A, Burlingham A, Cunningham R, Harper M, Jones AM, Violato M, Massey H, Strauss C. Challenging perspectives; understanding the barriers to engaging in an outdoor swimming feasibility randomised controlled trial. Health Place. 2024 Nov;90:103312. doi: 10.1016/j.healthplace.2024.103312.

38. Sheehan DV, Lecrubier Y, Sheehan KH, Amorim P, Janavs J, Weiller E, Hergueta T, Baker R, Dunbar GC. The M.I.N.I.: development and validation. J Clin Psychiatry. 1998;59(Suppl 20):22–33.

39. Database of Instruments for Resource Use Measurement (DIRUM). Clinical Service Receipt Inventory (CSRI) [Internet]. Bangor: DIRUM; [updated 2011 Jul 2011, accessed 2025 May 20] Available from: https://www.dirum.org/instruments/details/44

40. Spitzer RL, Kroenke K, Williams JBW, Löwe B. A brief measure for assessing GAD: the GAD-7. Arch Intern Med. 2006 May 22;166(10):1092. doi:10.1001/archinte.166.10.1092

41. Löwe B, Decker O, Müller S, Brähler E, Schellberg D, Herzog W, Herzberg PY. Validation and standardization of the Generalized Anxiety Disorder Screener (GAD-7) in the general population. Med Care. 2008 Mar;46(3):266–74. doi: 10.1097/MLR.0b013e318160d093.

42. Gu J, Strauss C, Crane C, Barnhofer T, Karl A, Cavanagh K, Kuyken W. Examining the factor structure of the 39-item and 15-item versions of the Five Facet Mindfulness Questionnaire before and after mindfulness-based cognitive therapy for people with recurrent depression. Psychol Assess. 2016;28(7):791–802. doi:10.1037/pas0000263

43. Herdman M, Gudex C, Lloyd A, Janssen M, Kind P, Parkin D, Bonsel G, Badia X. Development and preliminary testing of the new five-level version of EQ-5D (EQ-5D-5L). Qual Life Res. 2011 Dec;20(10):1727–36. doi: 10.1007/s11136-011-9903-x

44. Keetharuth AD, Brazier J, Connell J, Bjorner JB, Carlton J, Taylor Buck E, Ricketts T, McKendrick K, Browne J, Croudace T, Barkham M. Recovering Quality of Life (ReQoL): a new generic self-reported outcome measure for use with people experiencing mental health difficulties. Br J Psychiatry. 2018 Jan;212(1):42–49. doi: 10.1192/bjp.2017.10.

45. Keetharuth AD, Rowen D, Bjorner JB, Brazier J. Estimating a Preference-Based Index for Mental Health From the Recovering Quality of Life Measure: Valuation of Recovering Quality of Life Utility Index. Value Health. 2021 Feb;24(2):281–290. doi: 10.1016/j.jval.2020.10.012.

46. Patel A, Rendu A, Moran P, Leese M, Mann A, Knapp M. A comparison of two methods of collecting economic data in primary care. Fam Pract. 2005 Jun;22(3):323–7. doi: 10.1093/fampra/cmi027.

47. Bouwmans C, Krol M, Severens H, Koopmanschap M, Brouwer W, Hakkaart-van Roijen L. The iMTA Productivity Cost Questionnaire: A Standardized Instrument for Measuring and Valuing Health-Related Productivity Losses. Value Health. 2015 Sep;18(6):753–8. doi: 10.1016/j.jval.2015.05.009.

48. Kounali D, Button KS, Lewis G, Gilbody S, Kessler D, Araya R, Duffy L, Lanham P, Peters TJ, Wiles N, Lewis G. How much change is enough? Evidence from a longitudinal study on depression in UK primary care. Psychol Med. 2022 Jul;52(10):1875–1882. doi: 10.1017/S0033291720003700.

49. Batistatou E, Roberts C, Roberts S. Sample Size and Power Calculations for Trials and Quasi-Experimental Studies with Clustering. Stata J. 2014;14(1):159–75. doi:10.1177/1536867X1401400111

50. Adams G, Gulliford MC, Ukoumunne OC, Eldridge S, Chinn S, Campbell MJ. Patterns of intra-cluster correlation from primary care research to inform study design and analysis. J Clin Epidemiol. 2004 Aug;57(8):785–94. doi: 10.1016/j.jclinepi.2003.12.013.

51. Machin D, Campbell M, Tan S. Sample Size Tables for Clinical Studies. 3rd ed. Wiley; 2009.

52. Thorn JC, Davies CF, Brookes ST, Noble SM, Dritsaki M, Gray E, Hughes DA, Mihaylova B, Petrou S, Ridyard C, Sach T, Wilson ECF, Wordsworth S, Hollingworth W. Content of Health Economics Analysis Plans (HEAPs) for Trial-Based Economic Evaluations: Expert Delphi Consensus Survey. Value Health. 2021 Apr;24(4):539–547. doi: 10.1016/j.jval.2020.10.002.

53. Brueton V, Stenning SP, Stevenson F, Tierney J, Rait G. Best practice guidance for the use of strategies to improve retention in randomized trials developed from two consensus workshops. J Clin Epidemiol. 2017 Aug;88:122–132. doi: 10.1016/j.jclinepi.2017.05.010.

54. Grant S, Mayo-Wilson E, Montgomery P, Macdonald G, Michie S, Hopewell S, Moher D, on behalf of the CONSORT-SPI Group. CONSORT-SPI 2018 Explanation and Elaboration: guidance for reporting social and psychological intervention trials. Trials. 2018 Jul 31;19(1):406. doi: 10.1186/s13063-018-2735-z.

55. Montgomery P, Grant S, Mayo-Wilson E, Macdonald G, Michie S, Hopewell S, Moher D; CONSORT-SPI Group. Reporting randomised trials of social and psychological interventions: the CONSORT-SPI 2018 Extension. Trials. 2018 Jul 31;19(1):407. doi: 10.1186/s13063-018-2733-1.

56. National Institute for Health and Care Excellence. Health Technology Evaluation: The Manual [Internet]. Manchester: NICE; 2025 [updated 2022 Jan 31; accessed 2025 May 20]. Available from: www.nice.org.uk/process/pmg36

57. Husereau D, Drummond M, Augustovski F, de Bekker-Grob E, Briggs AH, Carswell C, Caulley L, Chaiyakunapruk N, Greenberg D, Loder E, Mauskopf J, Mullins CD, Petrou S, Pwu RF, Staniszewska S; CHEERS 2022 ISPOR Good Research Practices Task Force. Consolidated Health Economic Evaluation Reporting Standards 2022 (CHEERS 2022) statement: updated reporting guidance for health economic evaluations. BMC Med. 2022 Jan 12;20(1):23. doi: 10.1186/s12916-021-02204-0.

58. Faria R, Gomes M, Epstein D, White IR. A guide to handling missing data in cost-effectiveness analysis conducted within randomised controlled trials. Pharmacoeconomics. 2014 Dec;32(12):1157–70. doi: 10.1007/s40273-014-0193-3.

59. Leurent B, Gomes M, Carpenter J. Comment on: Sensitivity Analysis for Not-at-Random Missing Data in Trial-Based Cost-Effectiveness Analysis: A Tutorial Pharmacoeconomics. 2018;36(10):1297. doi:10.1007/s40273-018-0700-z

60. Hernández Alava M, Pudney S, Wailoo A. Estimating the Relationship Between EQ-5D-5L and EQ-5D-3L: Results from a UK Population Study. Pharmacoeconomics. 2023 Feb;41(2):199–207. doi: 10.1007/s40273-022-01218-7.

61. Personal Social Services Research Unit. Unit Costs of Health and Social Care Programme (2022–27) [Internet]. Canterbury: PSSRU; 2024 [accessed 2025 May 11] Available from: https://www.pssru.ac.uk/unitcostsreport/

62. National Institute for Health and Care Excellence. British National Formulary [Internet]. London: BMJ; 2025. [updated 2025 Apr; accessed 2025 May 11]. Available from: https://bnf.nice.org.uk/

63. NHS Business Services Authority. Prescription Cost Analysis – England [Internet]. Newcastle: NHS Business Service Authority; 2024. [updated 2024 Jun 06; accessed 11 May 2025]. Available from: https://www.nhsbsa.nhs.uk/statistical-collections/prescription-cost-analysis-england

64. Fenwick E, Marshall DA, Levy AR, Nichol G. Using and interpreting cost-effectiveness acceptability curves: an example using data from a trial of management strategies for atrial fibrillation. BMC Health Serv Res. 2006 Apr 19;6:52. doi: 10.1186/1472-6963-6-52.

65. Braun V, Clarke V. Using thematic analysis in psychology. Qual Res Psychol. 2006;3(2):77–101. doi:10.1191/1478088706qp063oa.

66. Dunn G, Emsley R, Liu H, Landau S, Green J, White I, Pickles A. Evaluation and validation of social and psychological markers in randomised trials of complex interventions in mental health: a methodological research programme. Health Technol Assess. 2015 Nov;19(93):1–115, v–vi. doi: 10.3310/hta19930.

67. Troncoso P, Morales-Gomez A. Estimating the complier average causal effect via a latent class approach using gsem. Stata J. 2022 Jun 30;22(2):404–15. doi:10.1177/1536867X221106416

68. Strauss C, Massey H. SPIRIT (2025) checklist for OUTSIDE 2: Outdoor swimming as a nature-based intervention for depression [Internet]. University of Sussex; 2026 [updated 2026 Mar2; accessed 2026 Mar12]. Available from: 10.25377/sussex.31445188

69. Medical Research Council (UK). Guidelines for Management of Global Health Trials [Internet]. MRC; 2022 [updated 2022 Feb 02, accessed 2023 Aug 21]. Available from: https://www.ukri.org/wp-content/uploads/2021/08/20220202_Guidelines-for-Global-Health-Trials-2017-v5-final.pdf

