## Supplementary material for "OUTSIDE 2: Outdoor swimming as a nature-based intervention, offered in addition to usual care, compared to usual care alone, in people with depression: A study protocol for a randomised controlled trial and economic evaluation": Statistical Analysis Plan

### OUTSIDE II

### Statistical Analysis Plan

**Trial registration:** ISRCTN Number (24759023); Registered prospectively on 21.02.2024.
**SAP version**: V3.0 17.11.2025
**Protocol version**: V11.0 08.07.2025

| Persons contributing to the analysis plan | |
|---|---|
| **Name** | Heather Massey |
| **Position** | Chief Investigator |
| **Name** | Stephen Bremner |
| **Position** | CTU Statistician |
| **Name** | Anna-Marie Bibby-Jones |
| **Position** | Trial Statistician |

| Authorisation | |
|---|---|
| **Name** | Stephen Bremner |
| **Position** | CTU Statistician |
| **Signature** | 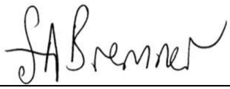 |
| **Date** | 17/11/2025 |
| **Name** | Heather Massey |
| **Position** | Chief Investigator |
| **Signature** | 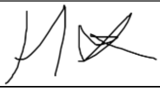 |
| **Date** | 17/11/2025 |

**Contents**

1. SAP revision history ... 4
2. Abbreviations ... 5
3. Introduction ... 7
3.1 Background and rationale ... 7
4. Study methods ... 7
4.1 Study design ... 7
4.2 Randomisation ... 7
4.3 Sample size ... 7
4.4 Framework ... 7
4.5 Statistical interim analyses and stopping guidance ... 7
4.5.1 Interim analyses ... 8
4.5.2 Early stopping guidelines ... 8
4.6 Timing of final analysis ... 8
4.7 Timing of outcome assessments ... 8
4.8 Blinding ... 8
5. Statistical principles ... 8
5.1 Confidence intervals and p values ... 8
5.2 Adherence and protocol deviations ... 8
5.3 Analysis populations ... 8
6. Study population ... 9
6.1 Screening data ... 9
6.2 Eligibility criteria ... 9
6.2.2 Exclusion criteria ... 9
6.3 Recruitment ... 10
6.4 Withdrawal/follow up ... 10
6.5 Baseline participant characteristics ... 10
7. Analysis ... 13
7.2 Outcome definitions ... 13
7.3 Analysis methods ... 13
7.4 Additional analyses ... 15
7.5 Harms ... 16
Summary of adverse events/reactions and serious adverse events/reactions by each arm. ... 16
7.6 Statistical software ... 16
8. References ... 17

#### 1. SAP revision history

| Version updated | Updated version number | Summary of changes | Author of changes | Date |
|---|---|---|---|---|
| V0.1 10.01.2024 | V0.2 13.02.2025 | Editing | Anna-Marie Bibby-Jones | 13/02/2025 |
| | V0.3 18.02.2025 | Clarified defn of follow-up; Added prognostic factors; added subgroups | Anna-Marie Bibby-Jones | 18/02/2025 |
| | V0.3 18.02.2025 | Added trial reg details | Gail Murphy | 18/02/2025 |
| | V1.0 22.05.2025 | Amendments following review | Stephen Bremner, Anna-Marie, Mara Violato, Clara Struass, Heather Massey | 22/05/2025 |
| | V2.0 26.09.2025 | Added sensitivity analysis for the 28 participants not meeting MINI criteria; Amended Subgroup analysis to be reduced from 5 to 3 following TSC advice. Clarified Ethnicity groups. | Anna-Marie Bibby-Jones | 26/09/2025 |
| V2 26.09.2025 | V3 17.11.2025 | Added Blinding statement | Anna-Marie Bibby-Jones | 17.11.2025 |

#### 2. Abbreviations

| | |
|---|---|
| AE | Adverse Event |
| AR | Adverse Reaction |
| CI | Chief Investigator |
| CRF | Case Report Form |
| CSRI | Client Service Receipt Inventory |
| CTU | Clinical Trials Unit |
| DMEC | Data Monitoring and Ethics Committee |
| FFMQ-15 | Five-Facet Mindfulness Questionnaire |
| GAD-7 | General Anxiety Disorder-7 |
| GCP | Good Clinical Practice |
| GP | General Practitioner |
| IAPT | Increasing Access to Psychological Therapies |
| ICC | Intracluster correlation coefficient |
| ICS | Integrated Care Systems |
| ISF | Investigator Site File (This forms part of the TMF) |
| ISRCTN | International Standard Randomised Controlled Trial Number |
| LEAP | Lived Experience Advisory Group |
| NHS R&D | National Health Service Research & Development |
| ONS | Office for National Statistics |
| PCN | Primary Care Networks |
| PHQ-9 | Patient Health Questionnaire-9 |
| PPI | Patient and Public Involvement |
| PI | Principal Investigator |
| PIC | Participant Identification Centre |
| PIS | Participant Information Sheet |
| QA | Quality Assurance |
| QC | Quality Control |
| RCT | Randomised Controlled Trial |
| REC | Research Ethics Committee |
| ReQoL-10 | Recovering Quality of life - 10 items |
| SAE | Serious Adverse Event |
| SAR | Serious Adverse Reaction |
| SDV | Source Data Verification |
| SOP | Standard Operating Procedure |
| SSI | Site Specific Information |
| SUSAR | Suspected Unexpected Serious Adverse Reaction |

| | |
|---|---|
| TMF | Trial Master File |
| TSC | Trial Steering Committee |
| SMG | Study Management Group |

#### 3. Introduction

##### 3.1 Background and rationale

Between March and June 2020, the number of adults experiencing moderate to severe depression in the UK was reported to be nearly 1 in 5. The total cost of mental ill health in England was estimated at £105 billion per year in 2016. In 2018, 70.9 million prescriptions for antidepressants were issued. The literature also indicates that 10-20% of individuals diagnosed with depression go on to have chronic depression and will have persistent symptoms lasting over 2 years. 50% can develop a recurrent depressive disorder, where they experience repeated episodes of depression throughout their life.

The number of people experiencing mental health problems is increasing. Meanwhile, pressure for NHS talking therapies is growing. Therefore, more consideration needs to be given to how preventive and community-asset based approaches to mental health can be delivered effectively. There is growing evidence that access to green and blue spaces is linked to improving mental health. The popularity of open water swimming is growing. Many advocates are reporting transformative experiences and a greater connectedness to nature as well as benefits to their mental health.

The benefits of open water swimming have not yet been explored and tested robustly in an adequately powered randomized controlled trial (RCT). So far, we have conducted a feasibility study which has provided evidence of the potential to recruit to and retain in an RCT. It also demonstrated the safety record for conducting such a study. These promising findings provide a platform for a multi-site full-scale RCT to examine the clinical and cost-effectiveness of the swim course intervention.

Study objectives

To evaluate the clinical and cost effectiveness of OUTSIDE of an 8-session introductory outdoor swimming course in addition to usual care in adults with symptoms of depression.

#### 4. Study methods

##### 4.1 Study design

Pragmatic study, two parallel arms, superiority RCT comparing the new intervention (Outdoor Swimming Course) and usual care and usual care alone (UC);

##### 4.2 Randomisation

1:1, Stratified by site in permuted blocks of randomly varying length, using REDCap.

##### 4.3 Sample size

480 participants i.e. 240 per arm. 280 participants recruited in year 1 and 200 participants in year 2. See protocol for full details of the calculation.

##### 4.4 Framework

This is a superiority RCT. A hypothesis test will be used to test the superiority of being in the Outdoor Swimming Course arm compared to usual care alone.

##### 4.5 Statistical interim analyses and stopping guidance

##### 4.5.1 Interim analyses

None

##### 4.5.2 Early stopping guidelines

The study may be stopped at the request of the DMEC if they consider it unsafe to continue based on the findings presented in any interim analysis requested.

##### 4.6 Timing of final analysis

All at once starting in February 2026

##### 4.7 Timing of outcome assessments

baseline (pre-randomisation – Time 0), 12 weeks post randomisation (post intervention - Time 1) and 38 weeks post randomisation (follow-up – Time 2)

##### 4.8 Blinding

Blinding of the trial statistician in clinical trials risk assessment tool (BRAT) (Partlett) assessment tool was used to document decision making process for this study. Following discussion with the study team, this was amended to both Statisticians remaining blind until the point of database lock. After that point they will both be unblinded.

#### 5. Statistical principles

##### 5.1 Confidence intervals and p values

95% confidence intervals will be presented for all estimated unstandardised treatment effect estimates for clinical outcomes.

Tests will be significant at the 5% alpha level.

No adjustment for multiplicity because a primary has been stated and all other outcomes are secondary.

##### 5.2 Adherence and protocol deviations

A summary (count & percentage) of participants within the Outdoor Swimming Course arm who are intervention completers by attending at least four of eight swim sessions, will be provided.

A tabulation of protocol deviations will be provided.

##### 5.3 Analysis populations

All analyses will be conducted according to intention-to-treat principles i.e. the sample is composed of all participants with observed data for primary and secondary outcomes and will be analysed as per their randomisation arm allocation.

We will also conduct a complier average causal effect (CACE)(Dunn et al, 2005, 2015) analysis of the primary outcome defining compliers as participants who had at least 4 out of 8 Swimming sessions).

Participants who withdraw consent for their data to be included will be excluded from all analyses.

#### 6. Study population

##### 6.1 Screening data

Summary of screening data count and screen failures. Data to be presented as part of the CONSORT diagram (Schulz et al., 2010).

##### 6.2 Eligibility criteria

Decisions about inclusions and exclusions will be based on medical information provided by participants and reviewed by the trial medical team, but further information or tests ordered via the participant's General Practitioner may be required if indicated.

###### 6.2.1 Inclusion criteria

The inclusion criteria for this trial are:

1. Participants give fully informed consent to participate;
2. Clinically important symptoms of depression, as determined by the Patient Health Questionnaire 9 (PHQ-9) score of 8 and above
3. Meeting DSM 5 criteria for a current Major Depressive Episode on the Mini International Neuropsychiatric Interview [MINI 7.0.2] (32) (the MINI 7.0.2 is a short structured diagnostic interview with screening and full versions to assess for DSM-5 and ICD-10 psychiatric disorders);
4. Self-reported ability to swim a minimum distance in a heated pool for sea (50 m, 2 lengths of a normal swimming pool) and lake (25 m, 1 length of a normal swimming pool) sites, no swimming ability required for outdoor unheated swimming pool (lido);
5. Adult aged 18 years or older;
6. Able to understand spoken instructions in English or in a language spoken by the swim coaches that are recruited.

###### 6.2.2 Exclusion criteria

1. High risk of suicide (determined by asking the person reporting: (1) in the past month having taken active steps with the intention of dying, (2) greater than 10% probability of acting on suicidal thoughts in the next few days (3) or 50% or greater probability of acting on suicidal thoughts in the next 3 months, or (4) for those reporting 1-49% probability of acting on suicidal thoughts in the next 3 months, risk of acting on thoughts deemed high following review by a study psychiatrist);
2. Evidence of a current Psychotic Disorder, or experiencing acute psychotic symptoms in the last 6 months (ascertained by the MINI 7.0.2, further assessed by the study psychiatry team where necessary);

Self-reported using a health history questionnaire (see Appendix 1), followed up with the medical team if necessary, with either further discussion or further investigation:

3. History of serious cardiac abnormalities;
4. Respiratory conditions triggered by cold such as poorly controlled exercise-induced asthma (self-reported);
5. Cold-water urticaria;
6. Participants with a BMI of 17 or under;
7. Moderate to severe learning disability;
8. History of non-freezing cold injuries, if not mitigated (wearing boots/gloves).

Or,

9. Currently participating in another interventional research trial

##### 6.3 Recruitment

Information to be presented on the CONSORT flow diagram.

##### 6.4 Withdrawal/follow up

Level of withdrawal (withdrawal from the trial or withdrawal from the intervention), timing and reason information to be presented on the CONSORT flow diagram.

##### 6.5 Baseline participant characteristics

| **Participant Characteristic** | **Summary** | **Comments** |
|---|---|---|
| Age | mean (sd) min max | Don't publish min & max, use for checking the data. |
| Gender | count (%) | |
| Sex | count (%) | |
| Whether Transgender | | |
| Employment | count (%) | Working/not working |
| Sexual Orientation | count (%) | |
| Marital status | count (%) | In a relationship/not in a relationship |
| Ethnicity | count (%) | Ethnic minority broken down by broad categories.<br><br>**White** – White British, White Irish, Any other White background; **Mixed or Multiple ethnic groups** – Mixed: White |

| | | |
|---|---|---|
| | | and Asian, Mixed: White and Black African, Mixed: White and Black Caribbean, Another mixed background (please describe below); **Asian or Asian British** – Asian: Bangladeshi, Asian: Chinese, Asian: Indian, Asian: Pakistani, Another Asian background (please describe below); **Black, Black British, Caribbean or African** – Black or Black British: African, Black or Black British: Caribbean, Another Black or Black British background (please describe below); and **Other ethnic group** – Arab, Gypsy or Irish Traveller, Another traveller background (please describe below), I identify with another ethnic group (please describe below),<br><br>Prefer not to say. |
| Birth country | count (%) | |
| First language | count (%) | |
| Number_children_under_18 | count (%) | |
| Religion or belief | | |
| Number_children_over_18 | count (%) | |
| Number of Adults cared for | count (%) | |
| Education Qualification | count (%) | |
| Disability status | count (%) | |
| Housing status | count (%) | |
| Household Income | count (%) | |
| Level of swimming ability | count (%) | |
| Previous experience of swimming | Count (%) | None / some |
| Index of Multiple Deprivation (IMD) | | Use the latest release available |
| Previous Treatment for depression in last 3 months | Count (%) | No Treatment, Medication only, Talking Therapy only, both. This information will be provided by Heather. |
| Location | count (%) | 1. Chalkwell<br>2. Ilkley Lido<br>3. Jesus Green Lido<br>4. Jubilee Pools<br>5. Loddington farm lake<br>6. Lymington sea water baths |

| | | |
|---|---|---|
| | | 7. Notts County Country Park<br>8. Parliament Hill Lido<br>9. Rayrigg<br>10. Rocker Beach<br>11. Salford Quays<br>12. Sea Lanes<br>13. Taplow Lake<br>14. West Country Water Park<br>15. Falmouth<br>16. Bournemouth<br>17. Bishampton<br>18. Birkenhead<br>19. West Reservoir<br>20. Quay Lake Woking<br>21. Shoreham |

#### 7. Analysis

##### 7.2 Outcome definitions

| Type of data | Variable name | Details |
|---|---|---|
| Quantitative /Qualitative | 'Information About You' at the start of the baseline assessment | 13 items, including multiple-choice questions and free text boxes |
| Quantitative | PHQ-9 Symptoms of depression | 9 items<br>Likert scales rating from 0 (not at all) to 3 (nearly every day)<br>Score is total of 9 items.<br>Min 0, max 27 (Higher is worse) |
| Quantitative | GAD-7 Symptoms of anxiety | 7 items - Likert scale rating from 0 (not at all) to 10 (nearly every day).<br>Score is total of 9 items.<br>Min 0, max 21 (Higher is worse) |
| Quantitative | FFMQ-15 Mindfulness | 15 items - Likert scale rating from 0 (worst) to 10 (best).<br>Score is total of all 15 items<br>Subscale scores total of values in each subscale<br>Scoring information<br>Describe items: 2, 7R, 12.<br>Acting with awareness items: 3R, 8R, 13R.<br>Non-judging items: 4R, 9R, 14R.<br>Non-reactivity items: 5, 10, 15.<br>Reverse-phrased items are denoted by 'R' after the item number |

##### 7.3 Analysis methods

The flow of participants through the trial will be shown on a CONSORT flowchart (CONSORT SPI 2018 Extension) (Schulz KF et al, 2010, Montgomery et al 2018).

We will summarise swimming course attendance as number of sessions/8 attended and number (%) of completers (attending 4 out of 8 sessions) by swim site type and year.

Summary statistics will be presented for each time point (T0, T1, T2) by randomised arm. Normally distributed variables will be described by their means and SDs, skewed continuous variables by their medians and interquartile ranges and categorical variables by the frequency and proportion in each category. Proportion of items missing and the proportion of participants that have a score for each outcome measure.

We will compare data completers to non-completers across demographics and baseline clinical outcomes at T1 and T2. Fisher's exact and chi-squared tests will be employed as appropriate.

The primary analysis will adhere to intention-to-Treat (ITT) principles where all participants included in the analysis will be as per their random allocation. The primary outcome will be analysed using a linear mixed effects model. Treatment arm (swimming + usual care or usual care alone), time (T1 or T2), and treatment arm × time interaction added as fixed effects; baseline MINI item A5 (at least one previous episode of depression ever (derived from the MINI Depression Module), and PHQ-9 and GAD-7 (selected *a priori*) scores will be added as covariates;  Site/swim group will be added as a random factor (random intercept) (Kahan & Morris, 2012), and effects will be estimated independently by treatment arm [using the Stata option *residuals(independent, by(treat))*], and random subject effects will be included to account for correlation between repeated measurements on individual participants. The site/swim group factor categories indicate the site and swim group which will simultaneously account for year. Restricted Maximum Likelihood will be applied using the *REML* option and a small sample adjustment for the degrees of freedom will be applied using Stata's *dfmethod(krogers)*. For the primary endpoint, the adjusted between-arm difference in mean PHQ-9, the corresponding 95% confidence interval, and associated p-value will be estimated at 12 weeks post randomisation. All p-values will be considered significant at the 5% alpha level using two-sided tests.

Standardised (Cohen’s d) between-group treatment effect sizes for each outcome will be calculated by dividing the between-arm unstandardised effect at T1 and T2 by the baseline pooled standard deviation.

In addition to the analyses for all clinical outcomes, the between-arm differences for the primary outcome will be assessed in the context of whether the minimal clinically important difference (MCID) is contained within the 95% confidence intervals where PHQ-9 scale of MCID is a 1.7 point reduction (Kounali et al, 2022).

Secondary outcomes (between-arm difference of PHQ-9 at T2 and all other outcome measures at T1 and T2) will be estimated using the same methods.

We will descriptively analyse the percentage of participants in the non-clinical range on the PHQ-9 score for depressive symptoms at T1 and T2 where PHQ-9 score =  0-9 [non-clinical], 10-14 [moderate], 15-19 [moderately severe], 20+ [severe].

We will report the number and proportion of Serious Adverse Events (SAEs) and Adverse Events (AEs) by trial arm and the number and proportion that are deemed by the independent clinical monitor to be study-related (i.e. Serious Adverse Reactions or SARs and Adverse Reactions ARs). For any SARs and ARs we will report a brief, non-identifying description of the event. We expect to be alerted to more SAEs and AEs in the intervention arm due to weekly contact with the swim coach.

Missing data will be addressed following the outcome scale's guidance, or using statistical techniques as appropriate for handling the missing data and then a sensitivity analysis will be conducted. Linear mixed models are effective for handling missing data under the assumption that data are Missing At Random (MAR). They accommodate unbalanced data and use all available data points to maintain statistical power.

**Alternative analyses**

If the model for the primary analysis will not converge then we will re-run the primary analysis model with site included as a fixed effect.

**Sensitivity analyses**

We will conduct a secondary complier average causal effect (CACE)(Dunn et al, 2015) analysis of the primary outcome to assess the impact of compliance (attending 4 or more sessions out of 8) on the primary outcome. For this analysis a two-stage instrumental variable regression approach will be used with treatment assignment as the instrumental variable and an adjustment for baseline PHQ-9 and any other covariates included in the primary analysis; robust standard errors will be used to account for within-site clustering. Compliance will be defined as i) a continuous measure of the number of swim sessions attended and then ii) a dichotomous measure indicating that four or more swim sessions were attended. We will use a Structural Equation Model (SEM) (Troncoso & Morales-Gómez, 2022).

Participants will have an assessment post intervention. The sensitivity analysis will look at those who completed this assessment within 4 weeks of post intervention.

The 28 ineligible participants randomised who did not meet MINI criteria who were excluded from the primary analysis will be included in the sensitivity analysis.

A sensitivity analysis will also be conducted if any statistical techniques are applied to address missing data by comparing the analysis of all available data.

**Hypothesis tests**

*Primary:*

Hypothesis 1: The 8-session outdoor swimming course offered in addition to usual care, in comparison with usual care only, will lead to greater reductions in **depressive symptom** severity from baseline to post randomisation (post-intervention).

*Secondary:*

Hypothesis 2: The 8-session outdoor swimming course offered in addition to usual care, in comparison with usual care only, will lead to greater reductions in **depressive symptom** severity from baseline to **38 weeks** post randomisation (follow-up).

Hypothesis 3: The 8-session outdoor swimming course offered in addition to usual care, in comparison with usual care only, will lead to greater reductions in **anxiety** symptom severity from baseline to **12 weeks** post randomisation and from baseline to **38 weeks** post-randomisation.

Hypothesis 4: The 8-session outdoor swimming course offered in addition to usual care, in comparison with usual care only, will lead to greater improvements in **mindfulness** from baseline to **12 weeks** post randomisation and from baseline to **38 weeks** post-randomisation.

Hypothesis 5: The 8-session outdoor swimming course offered in addition to usual care, in comparison with usual care only, will be **cost-effective** at **38 weeks** post-randomisation.

#### 7.4 Additional analyses

We will do subgroup analyses by Ethnicity, Gender, and type of swim location (e.g. lido, sea, lake) by adding the subgroup variable of interest plus an interaction term between the subgroup variable and treatment arm.

Summary data for between-arm differences will be provided for each category. 95% CIs and p-values will be provided for estimated treatment effects.

Findings should be treated with caution as subgroup-analyses tend to be underpowered and we may uncover spurious findings due to multiple analyses.

##### 7.5 Harms

Summary of adverse events/reactions and serious adverse events/reactions by each arm.

##### 7.6 Statistical software

Stata v19 (StataCorp, 2025) will be used for all analyses

#### 8. References

Dunn G, Maracy M, Tomenson B. Estimating treatment effects from randomized clinical trials with noncompliance and loss to follow-up: the role of instrumental variable methods. Stat Methods Med Res. 2005 Aug;14(4):369-95. doi: 10.1191/0962280205sm403oa. PMID: 16178138.

Dunn G, Emsley R, Liu H, Landau S, Green J, White I, Pickles A. Evaluation and validation of social and psychological markers in randomised trials of complex interventions in mental health: a methodological research programme. Health Technol Assess. 2015 Nov;19(93):1-115, v-vi. doi: 10.3310/hta19930. PMID: 26560448; PMCID: PMC4781463.

Kahan, B. C., & Morris, T. P. (2012). Analysis of multicentre trials with continuous outcomes: when and how should we account for centre effects? Statistics in Medicine, 32(7), 1136–1149. doi:10.1002/sim.5667

Kounali, D., Button, K., Lewis, G., Gilbody, S., Kessler, D., Araya, R., . . . Lewis, G. (2022). How much change is enough? Evidence from a longitudinal study on depression in UK primary care. Psychological Medicine, 52(10), 1875-1882. doi:10.1017/S0033291720003700

Montgomery P, Grant S, Mayo-Wilson E, Macdonald G, Michie S, Hopewell S, et al. Reporting randomised trials of social and psychological interventions: the CONSORT-SPI 2018 Extension. Trials. 2018 Jul 31;19(1):407

Partlett, C. Blinding of Trial Statisticians (BOTS) https://www.nctu.ac.uk/Our-Research/Methodology/Complete-Studies/BOTS.aspx [website accessed 17.11.25]

Schulz, KF., Altman, DG., Moher, D., for the CONSORT Group. CONSORT 2010 Statement: updated guidelines for reporting parallel group randomised trials. Trials. 2010;11:32. PMID: 20334632

StataCorp. 2025. Stata Statistical Software: Release 19. College Station, TX: StataCorp LLC.

Troncoso, P., Morales-Gómez, A. Estimating the complier average causal effect via a latent class approach using gsem. The Stata Journal: Promoting communications on statistics and Stata 2022. https://journals.sagepub.com/doi/10.1177/1536867X221106416?icid=int.sj-abstract.similar-articles

**Variables list from REDCap**

Data Dictionary

Copy%20of%20OUTSIDE2_DataDictionary
